# The de novo FAIRification process of a registry for vascular anomalies

**DOI:** 10.1101/2020.12.12.20245951

**Authors:** Karlijn H.J. Groenen, Annika Jacobsen, Martijn G. Kersloot, Bruna Vieira, Esther van Enckevort, Rajaram Kaliyaperumal, Derk L. Arts, Peter A.C. ‘t Hoen, Ronald Cornet, Marco Roos, Leo Schultze Kool

## Abstract

**Background:** Patient data registries that are FAIR - Findable, Accessible, Interoperable, and Reusable for humans and computers - facilitate research across multiple resources. This is particularly relevant to rare diseases, where data often are scarce and scattered. Specific research questions can be asked across FAIR rare disease registries and other FAIR resources without physically combining the data.

**Results:** We successfully developed and implemented a process of making a rare disease registry for vascular anomalies FAIR from its conception - *de novo*. Here, we describe the five phases of this process in detail: i) pre-FAIRification, ii) facilitating FAIRification, iii) data collection, iv) generating FAIR data in real-time, and v) using FAIR data. This includes the creation of an electronic case report form and a semantic data model of the “Set of Common Data Elements for Rare Disease Registration” released by the European Commission, and the technical implementation in the Electronic Data Capture (EDC) system for real-time, automatic data FAIRification. Further, we describe how we contribute to the four facets of FAIR, and how our FAIRification process can be reused by other registries.

**Conclusions:** In conclusion, a detailed de novo FAIRification process of a registry for vascular anomalies is described. To a large extent, the process may be reused by other rare disease registries, and we envision this work to be a substantial contribution to an ecosystem of FAIR rare disease resources.

## Background

Rare disease (RD) registries contain valuable information for improving diagnosis, treatment and event prevention [1][2]. Due to the low prevalence of RDs, using this information for research generally requires data from more than one registry. However, RD registries are distributed across the world and data from these registries are available in heterogeneous formats and multiple languages. As a consequence, optimizing the use of RD registries for research requires substantial efforts, and may be further complicated by legal constraints and the need for proper precautions for protecting the privacy of the sensitive data.

The FAIR data principles aim to enable efficient analysis of data across multiple sources through enhancing their Findability, Accessibility, Interoperability and Reusability for humans and computers [3]. Data that are FAIR at their source do not need to be physically combined for multi-source analysis. Multiple FAIR sources can be queried to answer research questions simultaneously in so-called federated queries. FAIR data are not open by definition. FAIR implies well-defined, transparent access conditions, which supports making data as open as possible and as closed as necessary. By applying the FAIR principles to RD registries (here referred to as the data collected from RD patients), analysis across multiple RD registries and other relevant FAIR data is made possible, even when access criteria differ per source.

The added value of the FAIR principles for RD research led to early acknowledgement by the RD community, and in 2017 the FAIR principles became a recognised resource by the International Rare Disease Research Consortium (IRDiRC) [4]. For example, since 2014, “Bring Your Own Data” workshops (BYODs) have been held to accelerate the adoption of the FAIR principles [5][6][7]. This includes a series of annually recurring BYODs in the RD domain. Over the years, the FAIRification process applied in BYODs has been explored and refined, and finally described step-by-step in a generic workflow [8]. An important step of this FAIRification process is to make data interoperable and machine-readable in a format that can be read and processed by computers. Machine-readable data is structured with the use of ontologies, which ensures that data may be more easily combined across RD registries, i.e. also if they are not in one database. IRDiRC has recognised ontologies to describe e.g. phenotypes (Human Phenotype Ontology - HPO [9]) and rare diseases (Orphanet Ontology for Rare Disease – ORDO [10]).

Another effort to further improve research across RD registries is the “Set of Common Data Elements for Rare Diseases Registration” (CDEs) released by the European Commission, Joint Research Centre [11]. The set consists of 16 data elements that are considered to be essential for research. Next to this, the European Commission has set up the European Rare Disease Registry Infrastructure (ERDRI) to facilitate findability of RD registries [12], a backbone for the European Reference Networks (ERNs) [13]. The ERNs are virtual networks at a European level, involving healthcare institutes recognized as expert centres for specific RDs. ERNs aim to facilitate discussion on complex, or rare diseases and conditions that require highly specialised treatment. Also, they aim to concentrate knowledge and resources. To that end, ERNs have been provided funding to set up a registry [14]. Minimum requirements include support for the CDEs, linking registries and making them interoperable. The European Joint Programme on Rare Diseases (EJP RD) further supports registries in implementing the FAIR principles. VASCERN is the ERN focusing on rare multisystemic vascular diseases [15]. VASCERN is subdivided into thematic working groups, one of which is the Vascular Anomalies working group, VASCA [16]. VASCA includes nine centres with individual databases and data collection processes.

Here, we describe how we set up a FAIR registry for vascular anomalies (hereafter referred to as the VASCA registry) in one of the VASCA centres, Radboud University Medical Center. The objectives were to 1) base our VASCA registry on the CDEs and the FAIR principles to enable it for analysis across RD registries, and 2) implement *de novo* FAIRification in our VASCA registry, where data are made automatically FAIR upon collection. By doing all the hands-on work for the FAIRification before data collection, data is made FAIR through entering them into an Electronic Data Capture (EDC) platform. This mitigates the need for repeated post-hoc FAIRification operations, thereby saving time and budget for the actual FAIRification of the data in the VASCA registry. To our knowledge, this is the first attempt to create a de novo FAIR RD registry, and may therefore serve as an example for (and be reused by) other registries.

## Results

We present a workflow for the de novo FAIRification process of the VASCA registry (**Figure 1**, see Methods). In the following sections, we describe how our approach contributes to each of the four facets of FAIR, and how it can be reused for other RD registries. For an automated assessment on how our approach contributes to each of the FAIR principles see [17].

**Figure 1.**
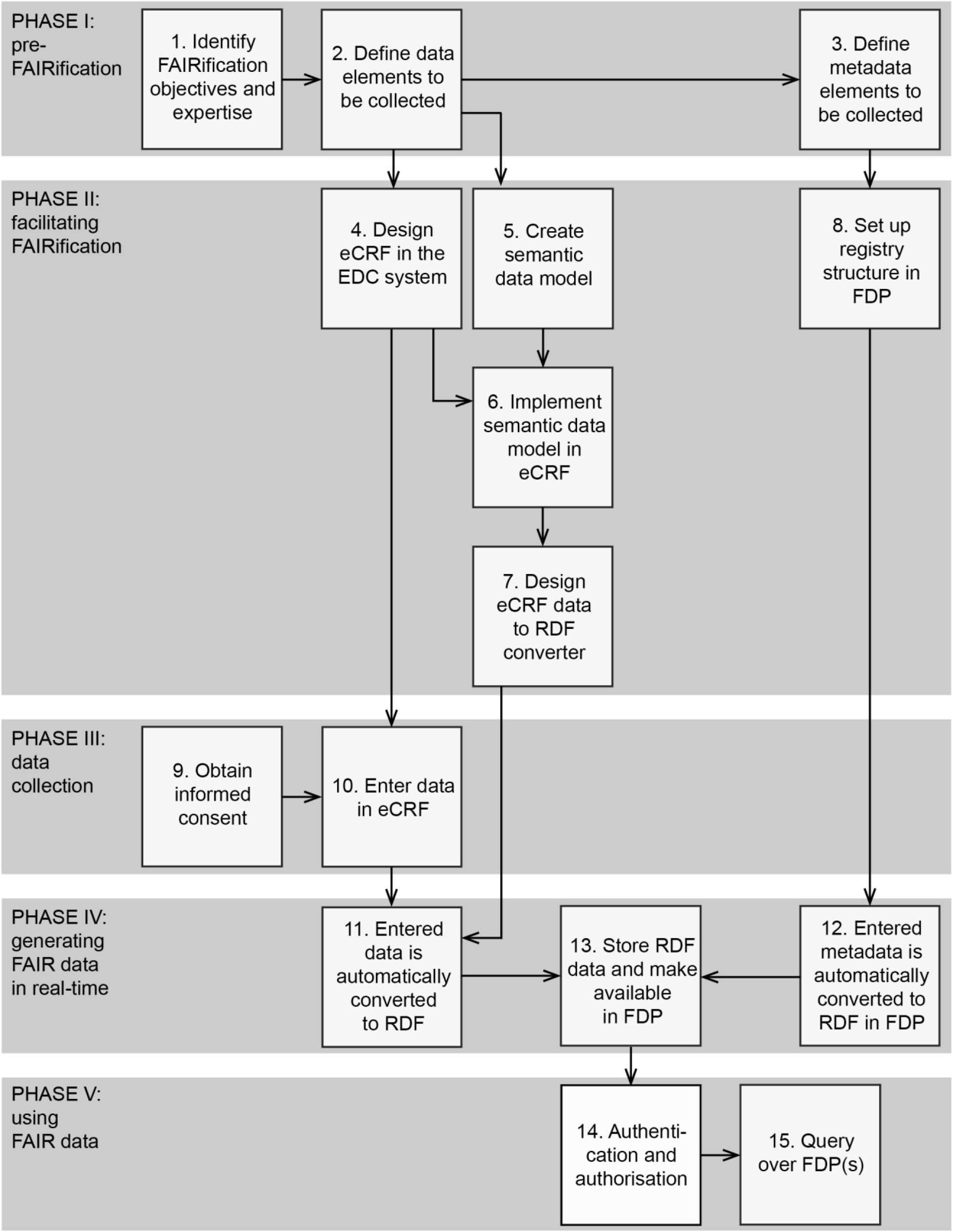
Workflow of the de novo FAIRification process of a registry for vascular anomalies. The workflow is divided into five “phases”: pre-FAIRification, facilitating FAIRification, data collection, generating FAIR data in real-time, and using FAIR data. The phases are further specified by “steps” indicating practical FAIRification tasks. Abbreviations: electronic Case Report Form (eCRF), Electronic Data Capture (EDC) system, Resource Description Framework (RDF; machine-readable language), FAIR Data Point (FDP).

**Figure 2.**
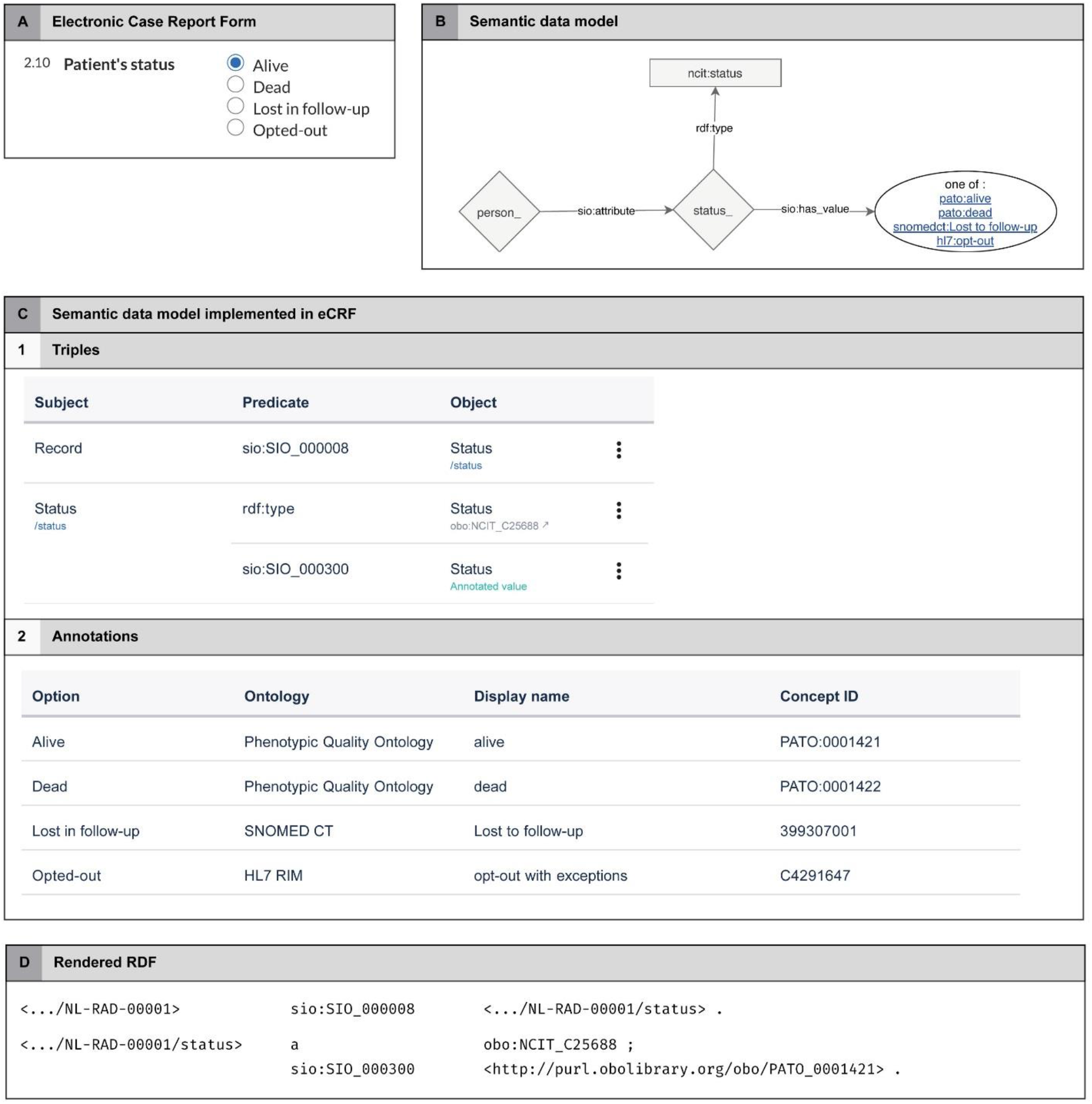
Schematic representation of the generation of RDF (d) based on eCRF data (a), a semantic model (b), and the implementation of this model in the eCRF (c). Abbreviations: Resource Description Framework (RDF; machine-readable language), electronic Case Report Form (eCRF)

### Contribution to the four facets of FAIR

#### Contribution to F - Findable

A description of the VASCA registry data (metadata) was provided to make it *findable* in searches for humans and computers. The metadata was structured using the Data Catalog Vocabulary (DCAT) standard and made machine-readable to enable searches by computer algorithms. Metadata for findability are for instance: database title (Registry of Vascular Anomalies), description (Databases of the ERNs vascular anomalies), and country (The Netherlands). The metadata was made available in a FAIR Data Point [18][19] and represented for humans in a visual interface and for computers in the Resource Description Framework (RDF) [20]: http://purl.org/castor/fdp/catalog/vasca.

VASCA registry metadata was also made findable for humans in the European Directory of Registries (ERDRI.dor) [21] under the namespace: ‘European Rare Vascular Anomalies Registry’. Metadata in ERDRI.dor include the medical areas involved, rare diseases registered, characterization of the registry, and affiliation to the ERN.

### Contribution to A - Accessible

The VASCA registry metadata in the FAIR Data Point is open and thereby *accessible* for all humans and computers. For privacy reasons, the registry patient data is not open, but access and querying may be granted (in compliance with the informed consent) after filing a request to the VASCA registry contact person as provided in the FAIR Data Point metadata (see Methods). Authentication (verifying the identity of the requestor) and authorization (verifying what the requestor is allowed to have access to) for gaining access to the patient data are arranged in the FAIR Data Point of the EDC system.

The VASCA registry can be seen in the ERDRI.dor overview, but the metadata related to the registry, including the metadata describing the institution/person responsible for the registry, is only available with a general login to the ERDRI.dor.

#### Contribution to I - Interoperable

The VASCA registry was made machine-readable and *interoperable* at a number of levels. First, the metadata was structured using the DCAT vocabulary following the FAIR Data Point specification. This contributes to machines being able to query the existence of the registry and its content descriptions. Second, the data collected in the eCRF were structured using an ontological model. The registry patient data were described using a semantic data model [22] constructed from terms and relations in commonly used ontologies (e.g. SNOMED CT and the IRDiRC recognised ontologies HPO and ORDO). Third, the VASCA registry was configured to collect data for the CDEs, and descriptions of these data elements were registered in the ERDRI Metadata Repository (ERDRI.mdr) [23] under the namespace: ‘European Rare Vascular Anomalies Registry’. The CDEs do not directly address any FAIR interoperability principles but do increase the compatibility of data in registries for certain analyses. Using an ontological model to define the meaning of these data elements ensures that we give access to a harmonized set of data elements and facilitate integration of CDEs from different registries, even across different ERNs. We note that without such an ontological model, computers cannot assess at source that common data elements are indeed common.

#### Contribution to R - Reusable

The VASCA registry data and metadata are sufficiently well-described to make them *reusable*. This means that they can be reproduced and repurposed (i.e. used for something else than their primary purpose). The metadata were published in the DCAT-based FAIR Data Point and made open using a CC0 license [24]. We included a number of elements in the metadata to support reusability. For example, the metadata contains a reference to an RDF ‘distribution’ of the data that can be queried in terms of the ontological model (see Methods, **Figure 3**). An example ontological query could be: “List all phenotypes reported for patients diagnosed with any type of vascular anomaly or angioma from VASCA FAIR Data Points in France, Germany, and The Netherlands”. We note that these queries can span multiple databases, because ontologies are not bound to a single dataset.

**Figure 3.**
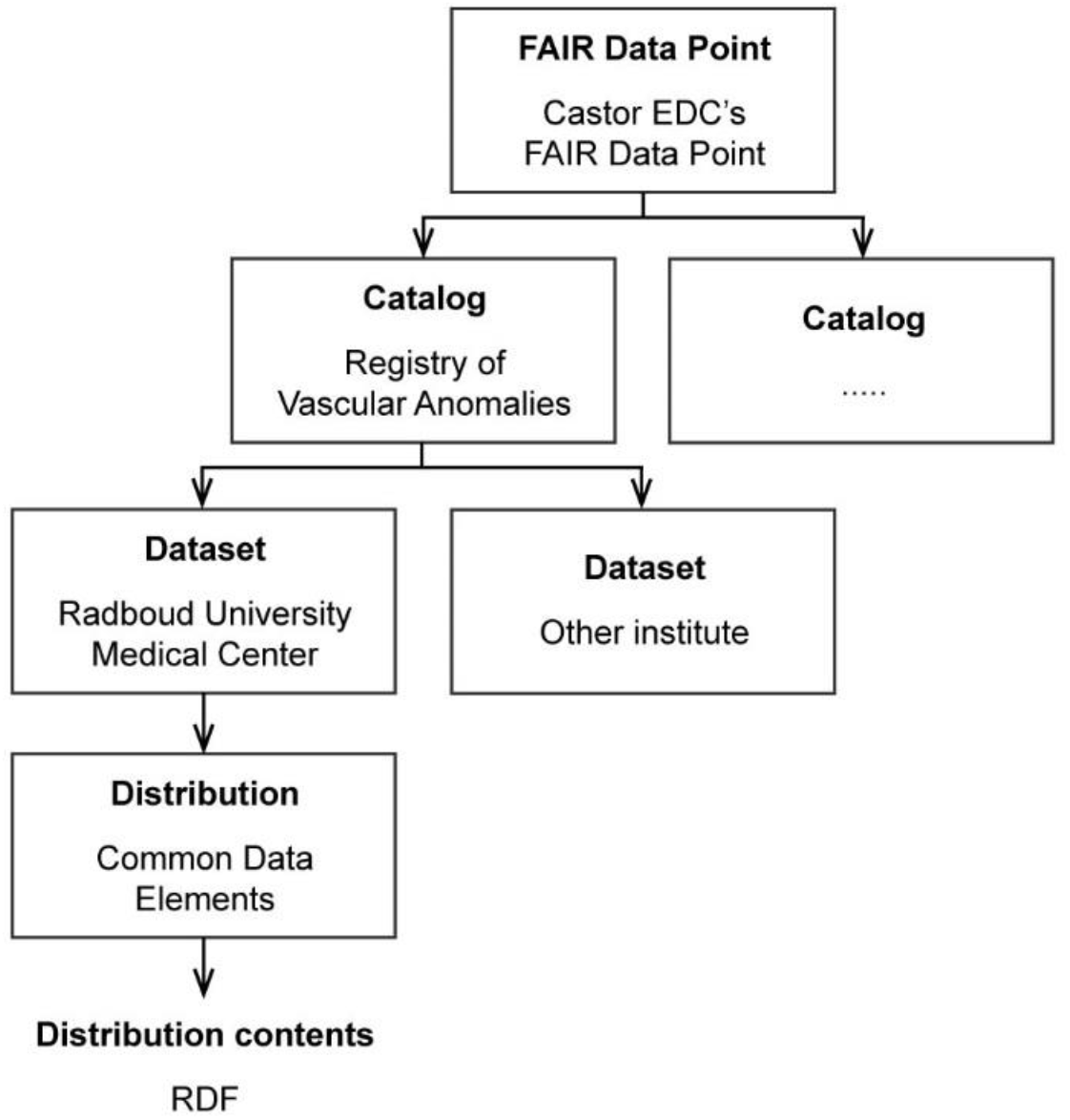
Metadata layers for the Registry of Vascular Anomalies (VASCA) in the Castor EDC’s FAIR Data Point. It consists of three layers: Catalog, Dataset and Distribution. The Distribution layer connects a machine-readable representation of the clinical data collected (in RDF; Resource Description Framework), only for authorized users.

The VASCA registry collects clinical data, which contains privacy sensitive data. By making it FAIR, the dataset is as closed as necessary and open as possible for other researchers (humans) and computers. Prior to collecting the data for the VASCA registry, informed consent was obtained. The informed consent form defines the options for reusing and exchanging data (see Methods). We complied to these options mainly by making the data available only to authorized users.

### Reusability of the de novo FAIRification process

Several aspects of the de novo FAIRification process of the VASCA registry have been made available and can be reused by ERNs for setting up their FAIR RD registries that collect the CDEs. The workflow (**Figure 1**) and expertise (**Table 1**) used in our FAIRification projects can be reused for organisation and preparation of other projects. Likewise, other aspects developed for our project that can be reused are our interpretations, semantic data model, and eCRF of the CDEs, FAIR implementations in Castor EDC, and structured metadata describing the VASCA registry. We interpreted the CDEs in order to define what data should be collected in the registry (Methods step 2). In our opinion the CDEs are multi-interpretable, hence, downstream implementations depend on these. We therefore properly defined and made our interpretations reusable for others in an extensive manual (available upon request).

**Table 1.**
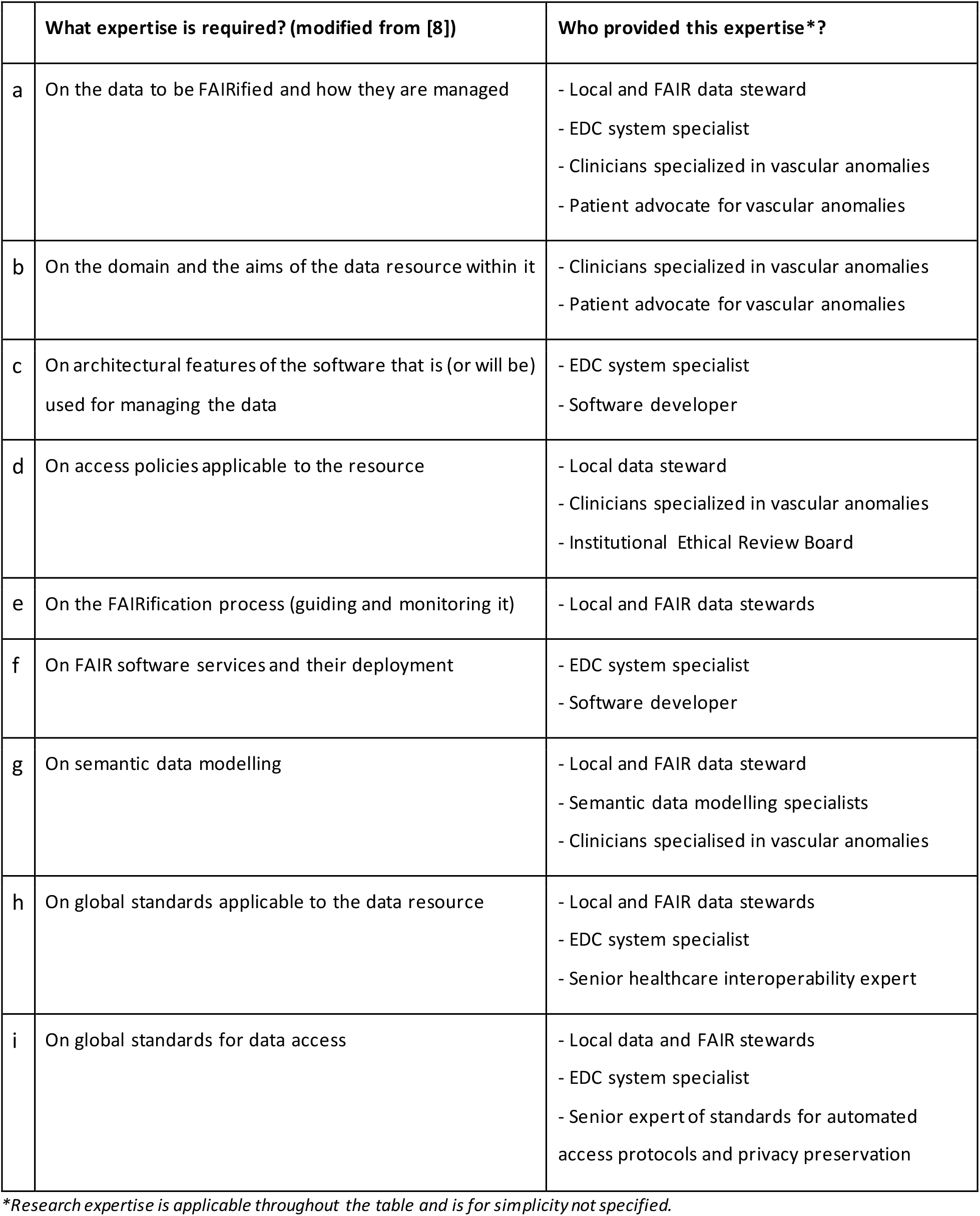
Expertise required for the FAIRification of a registry for vascular anomalies (VASCA). Research expertise is not specified as it is applicable throughout the table. The areas of expertise are inspired from previous FAIRification projects [8]. Abbreviations: Electronic Data Capture (EDC).

|  | What expertise is required? (modified from [8]) | Who provided this expertise*? |
| --- | --- | --- |
| a | On the data to be FAIRified and how they are managed | - Local and FAIR data steward<br>- EDC system specialist<br>- Clinicians specialized in vascular anomalies<br>- Patient advocate for vascular anomalies |
| b | On the domain and the aims of the data resource within it | - Clinicians specialized in vascular anomalies<br>- Patient advocate for vascular anomalies |
| c | On architectural features of the software that is (or will be) used for managing the data | - EDC system specialist<br>- Software developer |
| d | On access policies applicable to the resource | - Local data steward<br>- Clinicians specialized in vascular anomalies<br>- Institutional Ethical Review Board |
| e | On the FAIRification process (guiding and monitoring it) | - Local and FAIR data stewards |
| f | On FAIR software services and their deployment | - EDC system specialist<br>- Software developer |
| g | On semantic data modelling | - Local and FAIR data steward<br>- Semantic data modelling specialists<br>- Clinicians specialised in vascular anomalies |
| h | On global standards applicable to the data resource | - Local and FAIR data stewards<br>- EDC system specialist<br>- Senior healthcare interoperability expert |
| i | On global standards for data access | - Local data and FAIR stewards<br>- EDC system specialist<br>- Senior expert of standards for automated access protocols and privacy preservation |

\*Research expertise is applicable throughout the table and is for simplicity not specified.

We created a semantic data model that describes the CDEs and their relation and made it available on GitHub [22] (Methods step 5). Efforts to further develop and maintain the model are taking place [25] (also see Discussion). The goal of sharing the model is twofold: 1) Reuse: other ERNs can directly implement the model and would only need to extend the model with elements not represented in the CDEs. 2) Improving interoperability: It is easier to perform analysis across datasets/registries if they use the same semantic model. In that case, no ontology mapping is needed.

This project was done in close collaboration with Castor EDC [26], an electronic data capture system. Castor EDC developed the technology to facilitate the de novo FAIRification of the VASCA registry (phase ii in **Figure 1**). The eCRF designed for the CDEs, including the technology to translate to machine-readable format are reusable (Methods steps 6 and 7). The eCRF can one-on-one be copied to a new database in the Castor EDC, to initiate a new ERN registry. Some ERN-specific adaptations may be necessary. For instance, diagnosis is registered using a drop-down menu focussing on vascular anomalies and should therefore be adjusted for an ERN with a different focus. The ontologies used from the CDE semantic data model are not limited to an area of disease. The developed eCRF data to RDF transformation tool (Methods step 7) is generic and can be reused by other registries and clinical trials, ensuring that new FAIRification projects can easily be set up within Castor EDC. Likewise, other registries can reuse the FAIR Data Point structure and query functionalities developed for the VASCA registry (Methods steps 8, 13, 14, 15). Furthermore, we have made our eCRF interoperable and reusable, as the codebook describing the eCRF templates containing the CDEs and the ontologies to annotate them is openly available in ART-DECOR [27]. Via the openly available iCRF Generator tool [28], the codebook can be directly implemented in in EDC systems such as Castor EDC, OpenClinica and REDcap.

Finally, structured metadata describing the VASCA registry on ERDRI.mdr and the FAIR Data Point, respectively, are reusable. Structured record level metadata of the CDEs were included in ERDRI.mdr (name and descriptions of data collected for the CDEs). Other registries can clone and reuse the VASCA ERDRI.mdr metadata if they are setting up a registry according to the CDEs.

## Discussion

In this project, we set up our VASCA registry based on the CDEs and successfully showed the feasibility of creating a FAIR registry, automatically and in real-time, that allows for research across registries. The step-by-step description provided in this paper, might help other ERNs setting up their own FAIR registries. In the following sections, we discuss the lessons we learned during the project and describe our ideas for future developments.

### Lessons learned

#### The Common Data Elements

The CDEs include seemingly simple elements that turned out to be multi-interpretable. As an example, ‘sex’ can be interpreted as both genotypic sex and declared sex. Or, the element ‘date of first contact with a specialised centre’ requires a clear definition of a specialised centre; should it be a Healthcare Provider (HCP) that is a full ERN member, or can it also be an expert unit not being part of the ERN yet? It is essential for the interoperability of a registry that the CDEs are interpreted uniformly by the clinicians and centres that collect the patient data. We recommend that all registries clearly document their interpretations of the CDEs, for instance in a manual such as the one created for our VASCA registry. Ideally, guidelines are provided on a European level, improving interoperability across registries.

Another issue regarding the CDEs is the discrepancy between data to be collected for the registry and data that is actually collected within the Electronic Health Record (EHR) in daily clinical practise. For example, the ORPHAcodes used to define the diagnosis are very extensive and include a hierarchy. In clinical practice, clinicians may not use ORPHAcodes to code diagnoses in a patient’s medical record, nor use these detailed categories. Another example is the CDE ‘disability’. The EU prescribes to operationalize the CDE ‘disability’ using the WHO Disability Assessment Schedule (WHODAS). WHODAS, however, is only validated for adults, whereas a significant part of patients suffering from rare diseases are children.

Furthermore, the CDEs form a static description, thereby not capturing changes in the patients’ situation over time (follow-up). The data collected for the CDEs only represent their situation at the moment of data capture, but for some CDEs changes over time are likely to happen. For example, the execution of (new) diagnostic tests in a specialised centre or starting (new) treatments might very well affect the outcome of the disability score. Also, over time, new test results might become available (e.g. genetic tests, imaging), affecting the diagnosis. It is currently unclear in what cases and within what timeframe to update the information for already included patients. To this end, advice and alignment on when to assess and update the CDE data is needed.

The 16 CDEs form the core of the registries, but based on discussions with clinicians across Europe, we concluded that clinicians want to extend the dataset with disease-specific elements that differ between registries. This is however something that affects the work required for FAIRification, as the semantic data model should be extended with these disease-specific elements. Guidelines are required for extending the core CDE model with disease-specific elements. Also, between ERNs and/or registries coordination on data modelling is required to ensure compatible solutions (see also next section).

#### The semantic data model of the Common Data Elements

We learned that selecting ontologies can be difficult, as this process depended on the interpretation of the CDEs. When a CDE is interpreted similarly in different projects, it is recommended that the same ontology is used, as this prevents the need for mapping. To this end, we recommend that a standard set of ontologies should be defined for ERN registries (in addition to HPO and ORDO) to enhance interoperability. When a CDE is interpreted differently in different projects, correct interpretation by FAIR should be facilitated: differences in interpretation are acceptable as long as these interpretations are explicit and represented in a machine-readable format.

In the current project, interpreting the CDEs and selecting the corresponding ontologies were to some degree performed separately and independently. As shown in **Table 1** different expertises were required for interpretation of the data elements (clinicians specialised in vascular anomalies and patient advocate for vascular anomalies) and generating a semantic data model (local and FAIR data steward, semantic data modelling specialist and clinicians specialised in vascular anomalies). To enhance efficiency and quality of the semantic data model, we recommend both expertise to be at the table when developing and discussing the semantic data model (at least in the conceptual modelling part).

We have learned during our FAIRification project that the semantic data model continued to evolve after we implemented it in the eCRF. Currently, the model is being further developed and optimized by ontology experts in EJP RD. Besides this, in future we foresee ongoing adjustments due to e.g. improvement of technologies, ontologies as well as changes to the CDEs themselves. The question is if, how, and to what extent this would affect the interoperability of datasets. Therefore, one should think of how the community should deal with the use of different models (versions). Researchers should be able to use different versions of the model. Therefore, mapping between versions is essential. We foresee different approaches to deal with this. One would be that the ‘owner’ of the registry adjusts to a new model or new version. Another that developed models or versions are made mappable to earlier versions i.e. EJP RD should provide the community with either mapping tools or mappable models when further optimizing the CDE-based semantic data model. We would argue that the latter approach would be preferred as it costs less effort of the end users. Particularly if many users make use of the same model, where in the first approach, they would all adjust to the model individually. This also leads to further complexities such as different versions of semantics models needed to be mapped to different versions of the eCRF. In both approaches, our de novo FAIRification framework implies less extra work when a model is changed compared to post-hoc FAIRification. The conversion into a machine-readable format is more or less automatic and would only require implementing the updated model in the eCRF (Methods step 6). In contrast, post-hoc FAIRification would require additional redoing the semi-manual conversion into a machine-readable format.

#### FAIR implementation in Castor EDC

Enabling de novo FAIRification in Castor EDC required developing the necessary technology from scratch. We first piloted the generation of machine-readable data (RDF) to test the integration between the data transformation tool and the EDC system. We experienced that developing a generic tool that can be used by a large number of registries and clinical studies, despite that it took more time than developing a smaller registry-specific tool, is very important for making FAIR data available for other registries and researchers as well.

In addition, we decided to implement authentication and authorization layers in the FAIR Data Point by reusing the authentication and authorization of the EDC system. This means that at the moment, researchers that do not have access to the database in the EDC system are not able to access the data through the FAIR Data Point.

#### Informed consent

Informed consent is usually required for collecting prospective patient data for scientific purposes. The European Commission has provided the ERNs with a standard patient information folder (PIF) and broad informed consent form (ICF). Our Institutional Ethical Review Board did not approve the PIF and ICF for scientific registries. Main reasons were that the information provided on data handling was too limited. Therefore, our Institutional Ethical Review Board requested us to redraw the PIFs and ICFs. This has several possible consequences. Not only do the different centres need to follow local guidelines, one also needs to make sure data exchange is facilitated in an easy way. Future collaborations and/or data exchange should explicitly be part of the PIFs and ICFs.

### Future developments

The rapid development of FAIR technologies and possibilities requires us to continuously improve our FAIRification workflow. We are currently working on several aspects, discussed below.

The European Patient Identity Management (EUPID) pseudonymization tool [29] is recommended by the European Commission and aims to ensure that different registries can be mapped on a patient-to-patient level. However, at the time of setting up the VASCA registry, EUPID was not up and running yet and, therefore, not implemented in the VASCA registry. In the meanwhile, several developments related to the use of EUPID have been taking place. At the moment, EUPID provides a webservice that registries can connect to, in order to request its services. EUPID offers a specification guide where the methodology needed to call their web service is described, requiring local implementations of such requirements. We have identified three current ways of requesting EUPID services: via ERDRI, via the EDC system, or via the EHR system. First, ERDRI contains a web portal where the patient’s first and last name and date of birth can be manually inserted (individually or batch process). Subsequently, ERDRI, which is directly connected to EUPID services, provides a response. This option requires clinicians or data managers to use additional software, next to the EDC and EHR. The second option, using EUPID services via the EDC system, requires that identifiable patient information leaves the EHR system and ends up in the EDC system. This may not be desirable because research registries should only contain deidentified data. A better option from the viewpoint of automation, security, privacy and efficacy, therefore, would be to call on EUPID webservices from the EHR system directly. This option requires an application that collects the identifiable patient information from the EHR system, encrypts it according to EUPID requirements, sends the encrypted data to EUPID web service and handles its responses. We are currently exploring the technical options to integrate EUPID in the EHR system.

As described in the methods, we mapped the International Society for the Study of Vascular Anomalies (ISSVA) terms to the ORPHAcodes. However, the ISSVA terms not present in ORDO lacked a unique identifier. To comply with the interoperability principles, we are currently transforming the ISSVA classification into an ontology (OWL format), keeping the structure and adding all possible concepts and terms mappings to HPO, ICD, SNOMED CT, ORDO and NCIT. This way, in case an ISSVA term is not present in other existing ontologies, it has a unique identifier.

Setting up a registry requires a good balance between the amount of information one would like to collect, and the amount clinicians are able to provide given the limited time they can spend for each patient. In the current registry, clinicians provide all information. We are currently looking into the possibilities for a patient-driven registry. In patient-driven registries, patients fill in (part of the) data themselves rather than the clinician. This way, we would be able to collect more data with less effort. This would additionally enhance the options for collecting longitudinal data (which is not covered by CDEs), for example on quality of life, medication intake or treatments, thereby allowing additional research questions to be answered.

In addition, reaching interoperability requires the use of ontologies during data collection. Currently, this means that the data from the EHR (both structured fields and notes made by clinicians) should be ‘converted’ into terms used in the ontologies. This is manual work and is heavily dependent on interpretation by the person who does the data entry in the EDC system. To further optimize and automate this process, we are currently exploring whether software tools that automatically map free text to ontologies can aid in this. Examples are Phenotips [30], Zooma [31], and SORTA [32][33].

The web-based query method in the EDC system can currently only be used to query data in one registry, but work is being done to support querying over multiple registries. This would allow for easier retrieval of relevant information from multiple registries. For further interoperability, we would require an interface that facilitates queries over multiple registries, independent of the EDC system used for construction of the registries.

Next steps will also include the development of human and machine-readable access conditions to the data and, subsequently, the implementation of a mechanism for requesting and granting access to the data.

## Conclusion

In conclusion, we successfully set up a workflow for de novo FAIRification of the CDEs for the registry of vascular anomalies. The methods and lessons learned in the different phases of the FAIRification process are described in detail. This may help other ERNs in setting up their FAIR registries.

Next steps are to extend the VASCA registry with disease-specific data elements and to set up this registry in the other VASCA institutes and VASCERN working groups. This will allow us to analyse data across multiple registries using federated queries and, thereby, to demonstrate the added value of making them FAIR.

## Methods

### Workflow of the de novo FAIRification process

The FAIRification process for the VASCA registry developed and implemented in this project (**Figure 1**) distinguishes itself from the previously described generic post-hoc FAIRification process [8] in that it is implemented in the EDC system to make the data FAIR de novo. The workflow of the de novo FAIRification process is divided into five phases: i) pre-FAIRification, ii) facilitating FAIRification, iii) data collection, iv) generating FAIR data in real-time, and v) using FAIR data. The phases are further divided into steps describing practical FAIRification tasks detailed below. Through phases i-iv, the VASCA registry data and metadata become machine-readable. In the two final phases, the FAIR VASCA registry is made accessible (under well-defined conditions) and used for research.

### Phase 1: Pre-FAIRification

Pre-FAIRification pertains to the preparatory work before the actual implementation. Here, an inventory was made of everything necessary for developing and implementing the FAIR VASCA registry in a de novo manner. This includes e.g., objectives, team requirements, and availability of resources (data, tools, budget).

#### Step 1 - Identify FAIRification objectives and expertise

First, the FAIRification objectives for the VASCA registry were identified based on current challenges in RD research (for details see Background). The objectives help to set a scope for the FAIRification work to be done and to plan the FAIRification process. In short, these were to 1) base our VASCA registry on the CDEs and the FAIR principles to enable it for analysis across RD registries, and 2) implement de novo FAIRification in our VASCA registry, where data are made automatically FAIR upon collection.

Second, the expertise required to achieve the objectives were identified. Conducting the FAIRification process requires a highly multidisciplinary team guided by FAIR data steward(s) [8]. The core of the VASCA FAIRification team consisted of a local data steward, an external FAIR data steward, and an EDC system specialist. Throughout the project, additional expertise was consulted, such as a clinician specialized in vascular anomalies, the Institutional Ethical Review Board, and FAIR software developers and researchers. A full overview of the different kinds of expertise and which part of the FAIRification process they contributed to can be found in **Table 1**. Note, in our project, the expertise in many areas were provided by the same person. Also, research expertise is applicable throughout the table and is for simplicity not specified. The areas of expertise have been learned from a previous project [8], and further advanced here.

#### Step 2 - Define the data elements to be collected

As defined in the FAIRification objective, the data collected in the VASCA registry were based on the CDEs. The CDE ‘Classification of functioning/disability’ was not added, because there were many uncertainties about its use (see Discussion). We formally defined what data to collect for each of the other CDEs by interpreting the meaning of the CDEs and how they relate to each other. This was captured in an extensive manual for data collection. This task was done by the core FAIRification team together with clinicians specialised in vascular anomalies and a patient advocate. This was an important preparatory step for designing the eCRF (step 4) and creating the semantic data model (step 5) in the ‘facilitating FAIRification’ phase. This also contributed to consistent data entry by data managers across institutes, step 10 in the ‘data collection’ phase (see **Figure 1**).

#### Step 3 - Define the metadata elements to be collected

This step entails identifying what metadata (description of data) should be collected (e.g., license, owner, contributions statements, and description of use conditions and access of data) to comply with the FAIR principles. The World Wide Web Consortium (W3C) Data Catalog Vocabulary (DCAT) [34] is the default standard to predefine and structure metadata elements in the FAIR Data Point specification [35]. We decided to make the VASCA registry findable and accessible (under well-defined conditions) in a FAIR Data Point. Metadata elements described in the FAIR Data Point specification were therefore collected.

Metadata elements for the VASCA registry were also collected for ERDRI (ERDRI.dor and ERDIR.mdr). ERDRI.dor (European Directory of Registries) is a catalogue of RD registries. It contains the metadata related to the registry and includes 38 attributes, out of which 23 are compulsory. ERDRI.mdr (ERDRI Metadata Repository) contains detailed information about each variable collected in the registry including data type, description, and list of permitted items.

### Phase II: Facilitating FAIRification

In the second phase, the technical implementation in the EDC system was done to facilitate the de novo FAIRification. This pertains to e.g., the eCRF, the semantic data model, and the FAIR Data Point.

#### Step 4 - Design the eCRF in the EDC system

The eCRF was designed to collect data for the CDEs (described in step 2) Castor EDC [26]. Several dependencies (e.g. only show ‘Date of death’ when the patient is deceased) and validations (e.g. validate if the entered Online Mendelian Inheritance in Man (OMIM) genetic disorder code follows the OMIM standard) were included in order to collect high-quality data (for a detailed technical description see [17]). To this end, we mostly worked with closed questions and/or drop-down menus and prevented entering free text as much as possible. An example from the eCRF is shown in **Figure 2a**. The eCRF template containing the CDEs and the ontologies to annotate them (see step 5) was described in a codebook, which was made openly available in ART-DECOR, a platform from Nictiz, the Dutch competence centre for electronic exchange of health and care information [27], and which can be directly implemented in Castor EDC or other EDC systems using the openly available iCRF Generator tool [28].

#### Step 5 - Create the semantic data model

We created a semantic data model for the European Commission’s recommended set of CDEs to be used for the VASCA registry. The model is openly available on github [22]. A part of the model is shown in **Figure 2b**. First, our interpretations of the CDEs from step 2 were used to draw a conceptual model (listing the main concepts and relationships between the CDEs). This was done in close collaboration between the core FAIRification team, semantic data modelling experts and clinicians specialised in vascular anomalies to ensure that the intended meaning was captured. Later, machine processable ontologies were selected to replace the concepts in the conceptual model. A consequence of this process is that the representation of the data in the semantic data model is a product of our experts’ interpretations of the CDEs. Currently, the semantic data model of the CDEs is assessed and further optimized by the RD community (see Discussion). The CDEs recommend diagnosis to be defined with the Orphanet Ontology (ORDO). However, the elements available in ORDO (ORPHAcodes) did not contain all terms in the ISSVA classification used clinically to classify diagnosis (i.e. some ISSVA terms were lacking in orpha). As a solution, we transformed the ISSVA classification into a machine-readable ontology and added mappings to ORDO. The more specific ISSVA terms not available in ORDO were mapped to more general available ORDO terms.

#### Step 6 - Implement the semantic data model in the eCRF

The semantic data model of the CDEs was entered into a data transformation tool developed and provided by the EDC system (described in [17]). An example of this implementation is shown in **Figure 2c**. The model was recreated in this tool and specific elements in the model that should be filled with eCRF data were marked as *value nodes*. Subsequently, we mapped the questions of the eCRF to these *value nodes*. The implemented semantic model can be reused in other databases that are built in the EDC system by creating a mapping between the *value nodes* and the questions on the eCRF.

#### Step 7 - Design an eCRF data to RDF converter

In this project, we developed a data transformation tool [17], which transforms the data collected by the eCRF to a machine-readable representation, the Resource Description Framework (RDF). An example of the rendered RDF is shown in **Figure 2d**. The RDF representation is used since we work with ontologized data in this project and RDF allows machine-readable representation of ontologies. The transformation tool gets eCRF data from the EDC provider using their Application Programming Interface (API) and uses this data to fill in the required value nodes from the semantic data model based on the previously mentioned mappings between the model and the eCRF (**Figure 2**).

#### Step 8 – Set up registry structure in the FAIR Data Point

The available semantic metadata model of the FAIR Data Point specification was used to describe the VASCA registry [35]. This model is based on the DCAT standard. The VASCA registry FAIR Data Point metadata is described in three layers: 1) catalog - a collection of datasets, 2) dataset - a representation of an individual dataset in the collection, and 3) distribution - a representation of an accessible form of a dataset, e.g., a downloadable file or a web service that gives access to the data for authorized users (**Figure 3**). A catalog may have multiple datasets, and a dataset may have multiple distributions. The VASCA registry described in this project (Registry of Vascular Anomalies - Radboud University Medical Center) is one of the datasets in the catalog (Registry of Vascular Anomalies). Other VASCA registries, from this or one of the other centres can also be described in this catalog. The semantic metadata model of the FAIR Data Point metadata specification was implemented in the Castor EDC FAIR Data Point. The metadata that describe the catalog, dataset, and distributions of the VASCA registry described in this project, is publicly available and licensed under the CC0 license.

### Phase III: Data collection

The third phase covers the actual collection of the clinical data including the process of obtaining informed consent.

#### Step 9 - Obtain informed consent

Our project was approved by the Institutional Ethical Review Board of the Radboud University Nijmegen Medical Centre. Informed consent is obtained for each patient. The ERN template for obtaining informed consent was not used. Instead, custom made patient information sheets and informed consent forms were applied (see Discussion). Informed consent includes the use of the patient’s medical data for the VASCA registry as well as using this data in combination with data collected in other European registries or databases.

#### Step 10 - Enter data in the eCRF

Currently, data collection for the VASCA registry is a manual process, where data from the EHR is entered into the eCRF. Here, the symptoms described by the clinicians in natural language were manually converted into terms from HPO by using the HPO website [9].

The CDEs are static data elements, meaning that they do not include (changes over) time. Therefore, most data were collected and entered in the eCRF at the first contact in our centre. However, not all information is available at this point. For example, diagnostic imaging and genetic tests may still need to be performed. The results from these tests may provide new insights, thereby affecting CDEs such as genetic diagnosis, phenotype and age at which diagnosis was made. To include missing data or update data elements, we built in a six-month check, conducted six months after inclusion. At this point, data collected for the CDEs may be updated based on (newly) available information in the EHR.

### Phase IV: Generating FAIR data in real-time

This phase entails the process of the actual de novo FAIRification of the VASCA registry. Here, the entered data and metadata are automatically converted into machine-readable representations. The machine-readable metadata constitutes the metadata in the FAIR Data Point, and the machine-readable data is stored in a triple store (i.e., a specialized database in which RDF (see **Figure 2d**) can be stored and against which queries can be made) and made available in the FAIR Data Point.

#### Step 11 - Entered data is automatically converted to RDF

When the data is entered in the eCRF, it is automatically and in real-time converted into a machine-readable format by the data transformation tool. Thus, the data is made machine-readable from the moment it is being collected: de novo FAIRification. This way, a periodic, manual conversion of the data into machine-readable language is not required, resulting in all data collected being available for reuse at any time. Also, updates in the semantic data model are automatically being translated into the RDF being generated. An additional benefit of this approach is that the clinical people tasked with clinical care and data entry do not need this knowledge to generate FAIR data.

#### Step 12 - Entered metadata is automatically converted to RDF in the FAIR Data Point

When the metadata is entered in the FAIR Data Point of the EDC system, it is represented in a human-readable format (a website, e.g. https://fdp.castoredc.com/fdp/catalog/vasca), and at the same time automatically converted into a machine-readable RDF representation, (e.g. the ttl format: https://fdp.castoredc.com/fdp/catalog/vasca?format=ttl).

#### Step 13 - Store RDF data and make it available in the FAIR data point

After transforming the data into a machine-readable format, it is stored in a triple store. The data transformation tool only generates (step 11) and stores (this step) a patient’s machine-readable eCRF data when data is being entered (collected or updated) in the EDC system (step 10). The URL (Uniform Resource Locator) providing access to the machine-readable data in the triple store is made available in the FAIR Data Point as an access URL in the Distribution layer (**Figure 3**).

### Phase V: Using FAIR data

The final phase describes how the FAIR VASCA data available in the FAIR Data Point can be accessed and queried for research.

#### Step 14 - Authentication and authorisation

The VASCA registry metadata in the FAIR Data Point is open and can be accessed by API calls. The actual registry patient data can only be accessed and queried by logging in with an authorized account of the EDC system (either downloading the RDF or querying the data using SPARQL). The process of providing access, authentication and authorization, is arranged in the FAIR Data Point. A specific contact person for the VASCA registry (a Data Catalog Vocabulary contact Point) to be contacted for requesting access to the data is provided in the metadata. Access is only granted if it complies with the informed consent (see step 9). The contact person has the authority to decide if access is granted or not. If access is granted an authorized account is provided.

#### Step 15 - Query over FAIR data point(s)

The machine-readable data is stored in a triple store and can therefore be queried using the query language SPARQL. Users with access to the data (described in step 14) can send queries to the triple store to access specific data. Query results can be displayed in multiple formats (e.g. JSON, XML, CSV or TSV). The SPARQL endpoint of Castor EDC can be queried by using external SPARQL clients or a web-based version that is available in Castor EDC’s FAIR Data Point. Currently, the web-based version can only query within a single database. Federated queries, therefore, need to be done with external clients. These (federated) queries allow researchers to ask questions to the FAIR VASCA registry and other FAIR RD registries and data resources (multi-source analysis of FAIR data).

## Data Availability

The manuscript describes what parts or the workflow are available for reuse and where information/ data can be found.

## Acknowledgements

We thank Maria Barea for her input as a patient advocate. We further acknowledge Pim Kamerling, Janet Vos and Loes van der Zanden for test reading our manuscript.

## List of abbreviations

API: Application Programming Interface
BOYDs: ‘Bring your own data’ workshops
CC0: Creative Common zero
CDEs: Set of Common Data Elements for Rare Disease Registrations
CSV: Comma Separated Value
DCAT: Data Catalog Vocabulary
eCRF: electronic Case Report Form
EDC: Electronic Data Capture (platform)
EHR: Electronic Health Record
EJP RD: European Joint Programme on Rare Diseases
ERDRI: European Rare Disease Registry Infrastructure
ERDRI.dor: European Rare Disease Registry Infrastructure – European Directory of Registries
ERDRI.mdr: European Rare Disease Registry Infrastructure – Metadata Repository
ERN: European Reference Network
EUPID: European Patient Identity Management
FAIR: Findable, Accessible, Interoperable, Reusable
FDP: FAIR Data Point
HPO: Human Phenotype Ontology
ICD: International Classification of Diseases
ICF: Informed Consent Form
iCRF Generator: Interoperable Case Report Forms Generator
IRDiRC: International Rare Disease Research Consortium
ISSVA: International Society for the Study of Vascular Anomalies
JSON: JavaScript Object Notation
NCIT: National Cancer Institute Thesaurus
ORDO: Orphanet Ontology for Rare Disease
OWL: Web Ontology Language
PIF: Patient Information Folder
RD: Rare disease
RDF: Resource Description Framework
SPARQL: SPARQL Protocol and RDF Query Language
SNOMED CT: Systematized Nomenclature of Medicine – Clinical Terms
TSV: Tab Separated Values
URL: Uniform Resource Locator
VASCA: Vascular Anomalies
VASCERN: European Reference Network on rare vascular diseases
W3C: World Wide Web Consortium
WHODAS: WHO Disability Assessment Schedule
XML: Extensible Markup Language

